# Distribution of 54 polygenic risk scores for common diseases in long lived individuals and their offspring

**DOI:** 10.1101/2021.06.23.21257874

**Authors:** Sophia Gunn, Michael Wainberg, Zeyuan Song, Stacy Andersen, Robert Boudreau, Mary F. Feitosa, Qihua Tan, May E. Montasser, Jeffrey R. O’Connell, Nathan Stitziel, Nathan Price, Thomas Perls, Nicholas J. Schork, Paola Sebastiani

## Abstract

A surprising and well-replicated result in genetic studies of human longevity is that centenarians appear to carry disease-associated variants in numbers similar to the general population. With the proliferation of large genome-wide association studies (GWAS) in recent years, investigators have turned to polygenic scores to leverage GWAS results into a measure of genetic risk that can better predict risk of disease than individual significant variants alone.

We selected 54 polygenic risk scores (PRSs) developed for a variety of outcomes and we calculated their values in individuals from the New England Centenarian Study (NECS, N = 4886) and the Long Life Family Study (LLFS, N = 4577). We compared the distribution of these PRSs among exceptionally long-lived individuals (ELLI), their offspring and controls and we also examined their predictive values, using t-tests and regression models adjusting for sex and principal components reflecting ancestral background of the individuals (PCs). In our analyses we controlled for multiple testing using a Bonferroni-adjusted threshold for 54 traits.

We found that only 4 of the 54 PRSs differed between ELLIs and controls in both cohorts. ELLIs had significantly lower mean PRSs for Alzheimer’s disease (AD), coronary artery disease (CAD) and systemic lupus than controls, suggesting genetic predisposition to extreme longevity may be mediated by reduced susceptibility to these traits. ELLIs also had significantly higher mean PRSs for improved cognitive function. In addition, the PRS for AD was associated with higher risk of dementia among controls but not ELLIs (p = 0.0004, 0.3 in NECS, p = 0.03, 0.93 in LLFS respectively). Interestingly, ELLIs did not have a larger number of homozygous risk genotypes for AD (T_NECS_ = -1.72, T_LLFs_ = 0.83) and CAD (T_NECS_ = -5.08, T_LLFs_ = -0.31) in both cohorts, but did have significantly larger number of homozygous protective genotypes than controls for the two traits (AD: T_NECS_ =3.10, T_LLFs_ = 2.2, CAD: T_NECS_ = 6.57, T_LLFs_ =2.36, respectively).

ELLIs have a similar burden of genetic disease risk as the general population for most traits, but have significantly lower genetic risk of AD, CAD, and lupus. The lack of association between AD PRS and dementia among ELLIs suggests that their genetic risk for AD is somehow buffered by protective genetic or environmental factors.

## Introduction

Exceptionally long-lived individuals (ELLIs) demonstrate lower risk and delayed onset of many age-related diseases, such as cancer, cardiovascular disease, and dementia [1-4]. Their siblings and offspring also display lower prevalence of all-cause mortality, cancer, diabetes, and cardiovascular disease, suggesting that their resilience against disease and death may be at least partly genetic [5, 6]. Thus, ELLIs and their families are examples of successful aging, and investigating the genetic factors that defer or impede a wide variety of diseases associated with mortality may lead to greater insight into how we can promote and extend healthy aging.

As posited by Beekman et al., the survival advantage of long-lived families attributed to lower risk for common diseases “may be explained by the presence of alleles protecting against diseases that contribute to population mortality or the absence of alleles promoting such diseases” [7]. Here we investigate this question by considering protective and risk variants together, instead of separately. It is possible that the combination of a lower burden of disease variants and the presence of protective variants that buffer their effects determine ELLIs’ resilience to morbidity and mortality, and by investigating them together we can better understand genetic risk for common diseases in ELLIs.

Spurred by the ever-increasing number and sample sizes of the data and results from large genome-wide association studies (GWAS), polygenic scores have emerged as a way to utilize GWAS summary statistics to assess an individual’s genetic risk for disease. In its simplest form, a polygenic score is a weighted sum of alleles where weights are determined by the effect estimates from GWAS. Intuitively, one may expect only variants that meet genome-wide significance threshold to be valuable for predicting disease. However, significant variants explain only a small fraction of the predicted genetic variance[8]. In alignment with the current understanding that complex traits tend to be highly polygenic, studies have shown that PRS prediction performance can increase substantially by including variants that fail to meet genome-wide significance [9]. These polygenic risk scores (PRSs) allow investigators to capture substantial genetic variance in a single measure.

Previous studies have found no significant difference in the number of known disease-associated variants between ELLIs and controls [7, 10-13]. However, these studies were limited in three ways: they were small, they did not consider effect sizes of genetic variants, and they were limited in scope to only genome-wide significant SNPs. More recent studies have considered differences in polygenic risk computed using the effect sizes of variants that met genome-wide significance between the offspring of ELLIs and controls in the UK Biobank, and found that the offspring of ELLIs had lower PRSs for cardiovascular traits [14]. An Australian study sought to replicate these findings using ELLIs themselves rather than their offspring and controls, but was unable to do so, possibly due to the small number of cases ELLIs (n = 294) [15].

In this analysis, we examine the distribution and predictive value of 54 previously described PRSs [16] in two different ELLI cohorts: the New England Centenarian Study and Long Life Family Study [6, 12]. We also consider the impact of homozygous genotypes of both risk and protective alleles by creating what we call *homozygous scores*, allowing us further insight into the genetic differences between ELLIs and controls. Finally, we assess the performance of the PRS for parental longevity. To our knowledge, this represents the largest assessment of polygenic risk scores in ELLIs to date.

## Methods

### Data Sources

#### New England Centenarian Study (NECS)

The NECS is a study of centenarians, their offspring, their offspring’s spouses, and unrelated controls whose parents died before reaching the median age of death of their birth year cohort [12]. Participants included in this analysis were enrolled between 1995 and 2018. The age of participants were carefully validated [17][17](Young, Desjardins, McLaughlin, Poulain, & Perls, 2010)[17][17,18] and participants were followed up annually [12]. Data collected include demographic information, medical history and medications, measures of cognitive and physical functions based on Blessed and Barthel score (centenarians only), and Telephone Interview for Cognitive Status (TICS) and Instrumental Activities of Daily Living (IADL) questionnaires (offspring and spouses only). Genome-wide genotype data were generated using Illumina SNP arrays 370, 670 and 1M, and imputed to the HRC panel using the Michigan Imputation Server. All subjects provided informed consent approved by the Boston University IRB.

#### Long Life Family Study (LLFS)

The LLFS is a family-based study of healthy aging and longevity that recruited approximately 550 families and 5,000 family members selected for familial longevity [6, 18]. Participants were enrolled at three American field centers (Boston, Pittsburgh and New York), and a European field center in Denmark. The age of the oldest participants was validated [19]. Data collected at enrollment and during annual follow ups include demographic information, medical history and medications, measurements of physical and cognitive functions. Genome-wide genotype data were generated using the 2.5K Illumina SNP arrays and imputed to the HRC panel using the Michigan Imputation Server. All subjects provided informed consent approved by the field centers’ IRBs. Data are available from dbGaP (study accession: phs000397.v1.p1).

#### Illumina Controls

This repository includes approximately 3500 samples used as controls of a variety of genome wide association studies. The data can be accessed using the protocol available from here (http://www.illumina.com/documents/icontroldb/document_purpose.pdf). We used this set of controls as referent population in study of longevity since we expect that only a small portion of them would become centenarians. Genome-wide genotype data were generated with a variety of Illumina SNP arrays and quality control was previously described in [20]. Genotype data were imputed to the HRC panel using the Michigan Imputation Server as in the other cohorts.

### Polygenic Risk Scores

We calculated polygenic risk scores (PRSs) based on public genome-wide summary statistics for 54 diseases (Supplementary Table 1), as described previously [16]. Briefly, we considered only variants from the summary statistics with p < 1 × 10^−3^, then pruned to variants with linkage disequilibrium (LD) r^2^ < 0.2. PRSs were scored on the NECS and LLFS individuals using the “--score” function from version 2.00 of the *plink* software package[21], by summing the dosages of the variants in the PRS weighted by GWAS effect size. Not all variants in the original PRS were available in our cohorts (Table S1), due to using an imputation quality threshold of 0.7 for both cohorts.

#### Homozygous Scores

We computed three types of scores to assess whether ELLIs and their offspring were more or less likely to be homozygous for the minor alleles in our PRSs than controls. First, we classified the variants in each trait’s PRS as risk-increasing or protective, based on the effect direction of the minor allele. We then created a simple count of how many protective minor alleles each individual was homozygous for, which we call the *protective homozygous count*. We also performed the same calculation for risk-increasing variants, which we call the *risk homozygous count*. These two scores allow us to count and individuals’ number of homozygous genotypes for protective and risk minor alleles. Finally, we scored the individuals using the “recessive” option to plink 2’s “--score” function, which only assigns genetic risk (both positive or negative depending on the set of variants used) to individuals with dosages greater than 1 for the coded allele of the PRS. We call this score a *homozygous PRS*. This score allows us to look at an individuals’ homozygous genotypes, weighted by the effect estimate of the minor allele.

### Statistical analyses

In the LLFS, we used three definitions of ELLIs based on the attained age at last contact: individuals were classified as ELLIs if they survived to an age reached by less than 10%, 5% or 1% individuals in their birth year and sex specific cohort. In the NECS, we defined centenarians and their nonagenarian siblings as ELLIs (see Table S2).

#### Distribution Analysis

To evaluate the mean difference in PRS between ELLIs and controls for each trait, we used a linear mixed model to predict the PRS by case status, adjusting for sex, the top six genotype principal components, with a random effect for family ID, using a Bonferroni-adjusted threshold of *P < 0*.*00092* (i.e. 0.05 divided by 54) to control for multiple testing. Since the clinical utility of PRS appear to lie at the upper quantiles of the score [9], we also tested whether there was a difference in proportion above the 90^th^ percentile in PRS between ELLIs and controls.

#### Association with Disease Risk

We used a mixed effect Cox proportional hazard regression model to predict incidence of Alzheimer’s disease and dementia, cardiovascular disease and myocardial infarction using the corresponding PRSs. Age was censored at last contact. We conducted two distinct analyses: one using the PRS as a continuous variable, and another using a dichotomous variable indicating whether an individual had an exceptionally high PRS, as defined as the 90% percentile among the roughly 3000 controls in the NECS. We ran the continuous PRS model in both NECS and LLFS. However, since we used NECS to define the threshold for an exceptionally high PRS, we conducted the analysis of the dichotomous PRS only in LLFS. All models were adjusted for sex and the top six genotype principal components, with a random effect for family ID to account for familial-genetic correlation. We analyzed the controls and ELLIs separately. In LLFS, we limited the analyses to ELLIs in the top 5%, since there were too few individuals who lived past the 1% threshold, the 10% threshold was not sufficiently conservative. We tested the proportional hazard assumption using the cox.zph() function from the R package “survival”, which compares the Schoenfeld residuals against transformed time.

#### Homozygous Scores

To further examine the genetic differences between ELLIs and controls, we considered whether ELLIs were more likely to have homozygous genotypes for protective alleles, and less likely to have homozygous genotypes for disease risk variants. For AD and CAD, we calculated the number of homozygote genotypes with protective alleles, called *protective homozygous count*, and risk alleles, called *risk homozygous count*, and also created a *homozygous PRS*, which weights each homozygous genotype by the effect estimate of the minor allele, in our two cohorts, and assessed the differences in distributions of these scores. We tested whether there was a mean difference in the *protective homozygous count, homozygous count*, and *homozygous PRS* using the same technique as before, using a linear mixed model to predict the risk scores by case status.

We next associated each of the three types of homozygous scores with incidence of the corresponding disease (either AD or CAD), using the same procedure as described in the previous section. Here, we control for multiple testing adjusted for three types of homozygous scores with a Bonferroni significance threshold, of p = 0.017 (i.e. 0.05 divided by 3).

#### Parental Longevity

To assess the PRS for parental extreme longevity, we used a GEE model with an exchangeable correlation structure to predict parental extreme longevity. In the NECS, we subset the data to offspring of ELLI with known age at death, and to controls. In LLFS, we subset the data to only the offspring generation, which includes both the children of ELLI and their siblings, and their spouses, the controls. We analyzed the effect of the continuous PRS in both the LLFS and the NECS, and the dichotomous variable indicating exceptionally high PRS only in the LLFS. Each model was adjusted for sex, the first six principal components of ancestry and an exchangeable correlation structure to account for family correlation.

## Results

### Baseline Characteristics

NECS participants included 1495 ELLIs (73% female), 487 offspring of ELLIs (63% female), and 2899 controls (59% female). LLFS participants included1270 individuals who lived past the 10% survival age (47% female), 1044 who lived past the 5% survival age (43% female), and 502 who lived past the 1% survival age (41% female). See Table S2.

### Distribution of PRS

Figure 1 summarizes the differences of 11 PRSs that were significantly different between ELLIs in NECS at the Bonferroni-adjusted significance threshold, while the full set of results for the 54 traits is in Table S2. Consistent with the lower risk for many aging-related diseases of ELLIs [2], the PRSs for Alzheimer’s disease (AD), atrial fibrillation, BMI, coronary artery disease (CAD), stroke, systemic lupus, and systolic blood pressure were significantly lower in ELLIs, while the PRSs for education attainment, and cognitive function were significantly higher. The PRSs for IBD and ulcerative colitis was significantly higher in ELLIs compared to controls. The figure also shows the results comparing ELLIs and controls in the LLFS for the same 11 traits, and the PRSs for AD, CAD, and systemic lupus were significantly lower in ELLIs than controls while the PRSs for educational attainment and cognitive function were significantly higher in ELLIs than controls independently of the threshold used to define extreme longevity. The distribution of the PRSs for the other traits showed similar trend but with more moderate level of significance.

**Figure 1.**
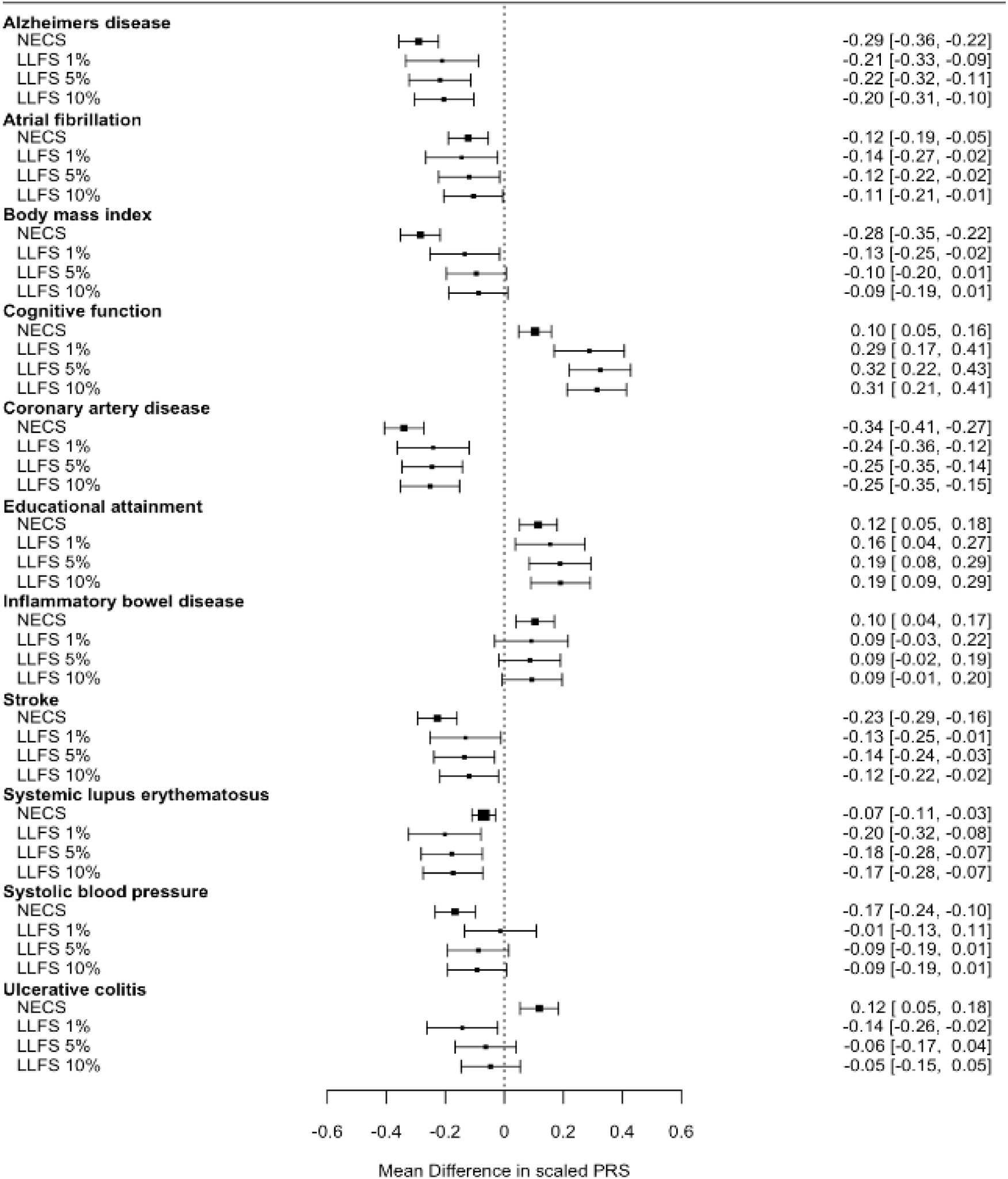
Estimated mean difference and 95% CI of scaled PRS between ELLI and controls for which we found a significant mean difference in NECS Extreme longevity was defined by survival thresholds depending on their sex in LLFS and centenarian status in NECS. Mean difference is in scaled PRS.

In the LLFS, we found a statistically significant mean difference in PRS between ELLIs and controls for an additional 12 traits (Figure 2). On average, ELLIs in LLFS had a lower PRS for height, birthweight and prostate cancer, and a higher PRS for amyotrophic lateral sclerosis (ALS), chronic kidney disease, atopic dermatitis, narcolepsy, psoriasis, gout, primary biliary cholangitis, and parental extreme longevity.

**Figure 2.**
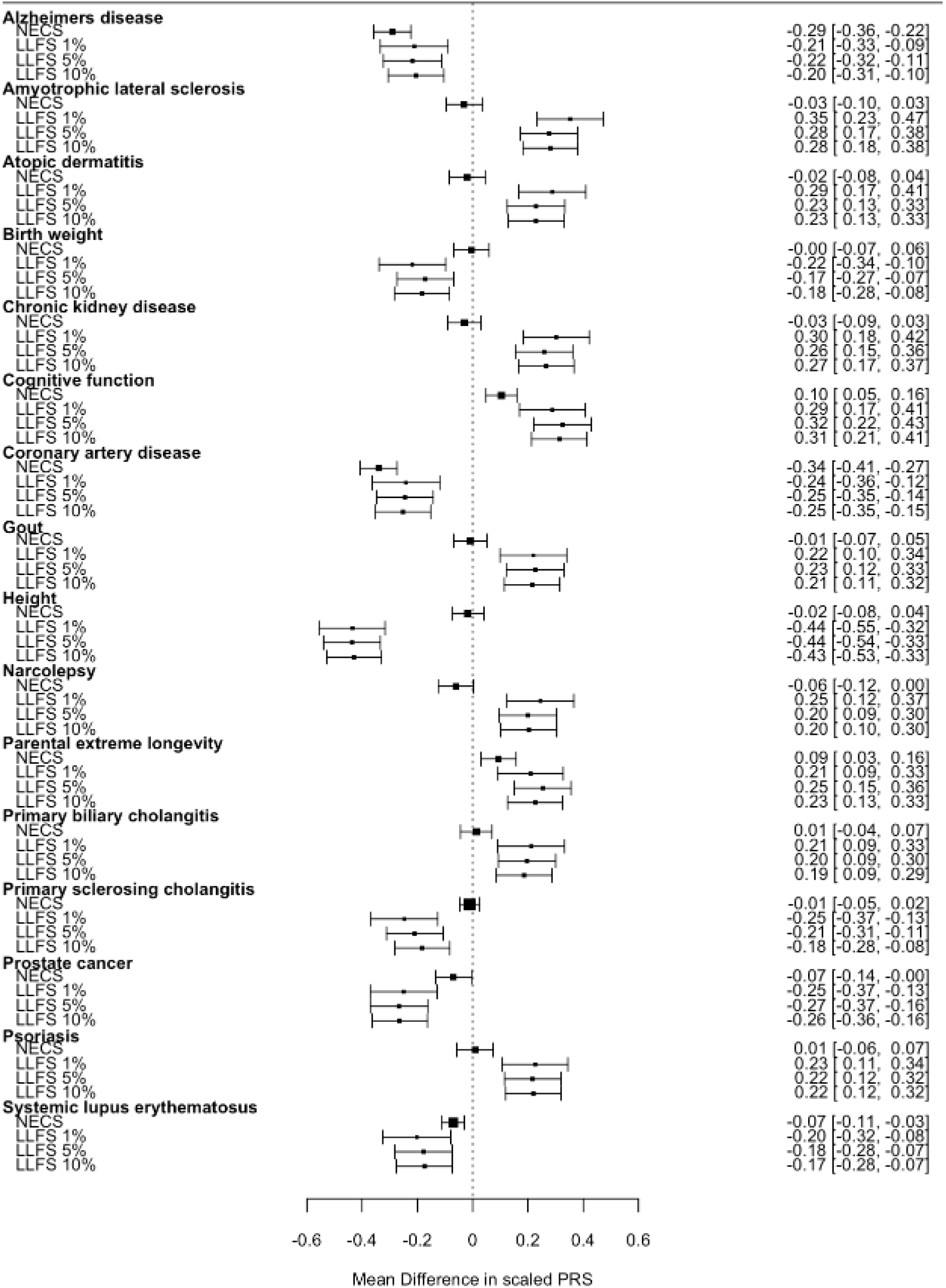
Estimated mean difference and 95% CI of scaled PRS between ELLI and controls for traits for which we found a significant mean difference in LLFS. Extreme longevity was defined by survival thresholds depending on their sex in LLFS and centenarian status in NECS. Mean difference is in scaled PRS.

Of the 11 PRSs associated in NECS and the 12 PRSs associated in LLFS, 4 PRSs were associated in both cohorts: Alzheimer’s disease (AD), coronary artery disease (CAD), lupus, and cognitive function. The distributions of these 4 PRSs demonstrates that at the tails there is clear separation between the ELLIs and controls (Figure 3). To formally test this observation, we computed the 90^th^ percentile of each of the four traits’ PRSs across the 3000 controls in NECS, then compared the proportion of LLFS controls with PRSs above this 90^th^ percentile threshold to the proportion of top 1% ELLIs in LLFS with PRSs above this threshold. For all four traits, the difference in proportion was statistically significant (see Figure 3 caption).

**Figure 3.**
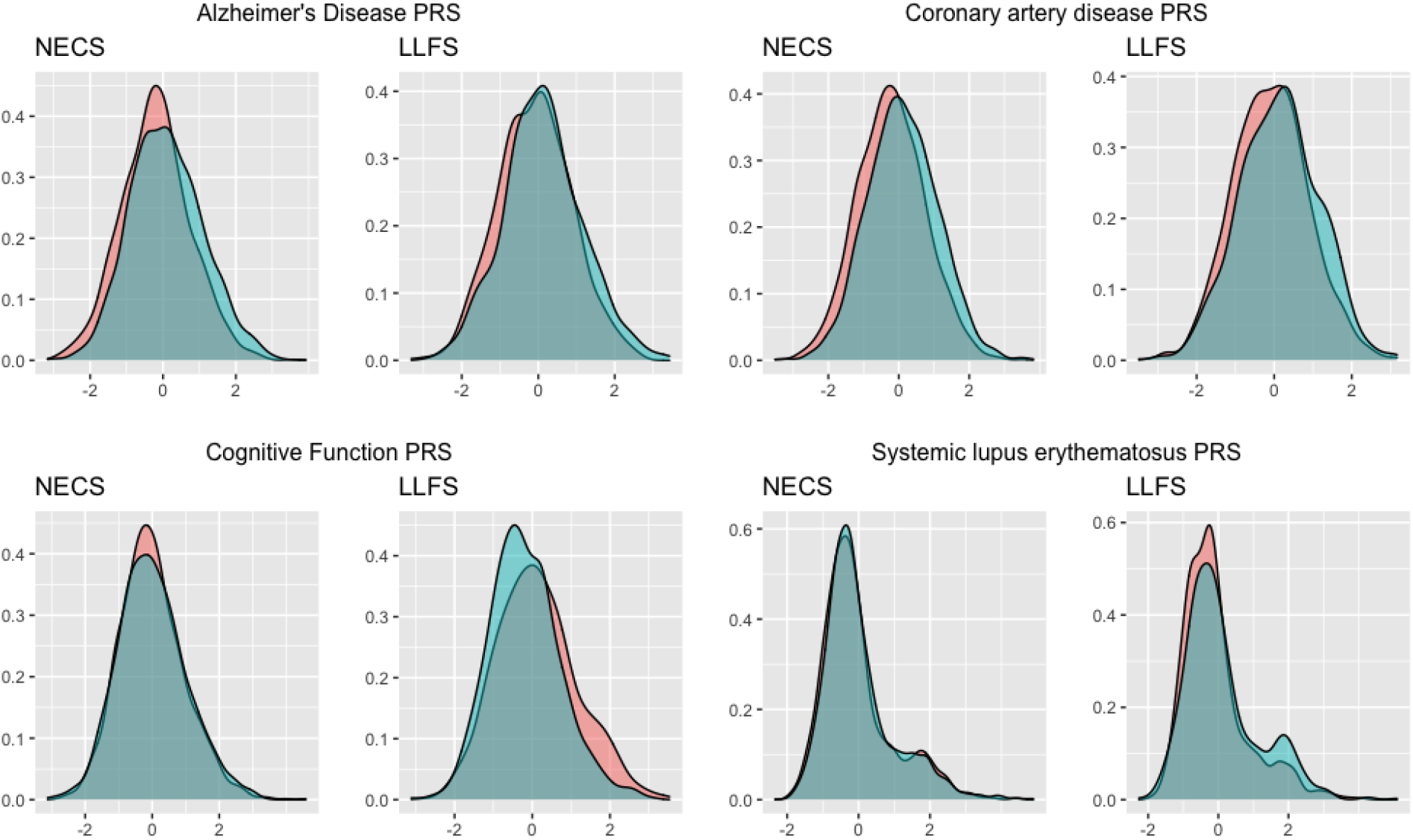
Distribution of PRS Density plots comparing scaled PRSs among ELLI (red) to controls (blue) for the four traits for which we found significant mean difference between the two groups at a Bonferroni adjusted significance threshold. ELLI status is defined in LLFS as 5% survival. Using the NECS controls to define a 90% percentile, for Alzheimer’s 10.2% of LLFS controls were past the threshold, compared to 5.2% of the ELLI (p = 6.15e-105) ; for CAD 13.4% of LLFS controls were past the threshold, compared to 7.8% of the ELLI (5.5e-89), for Cognitive function, 6.3% of LLFS controls were past the threshold, compared to 14.7% of the ELLI (p = 5.4e-126); and finally, for Lupus 15.2% of the LLFS controls were past the threshold, compared to 7.4% of the ELLI(p = 2.7e-80).

We also compared the distribution of PRSs between the offspring of ELLIs in LLFS and controls in the LLFS. This analysis showed that the offspring of the extremely long lived had higher polygenic risk for ALS, cognitive function, and parental extreme longevity, and lower polygenic risk for CAD and height (Figure 4).

**Figure 4.**
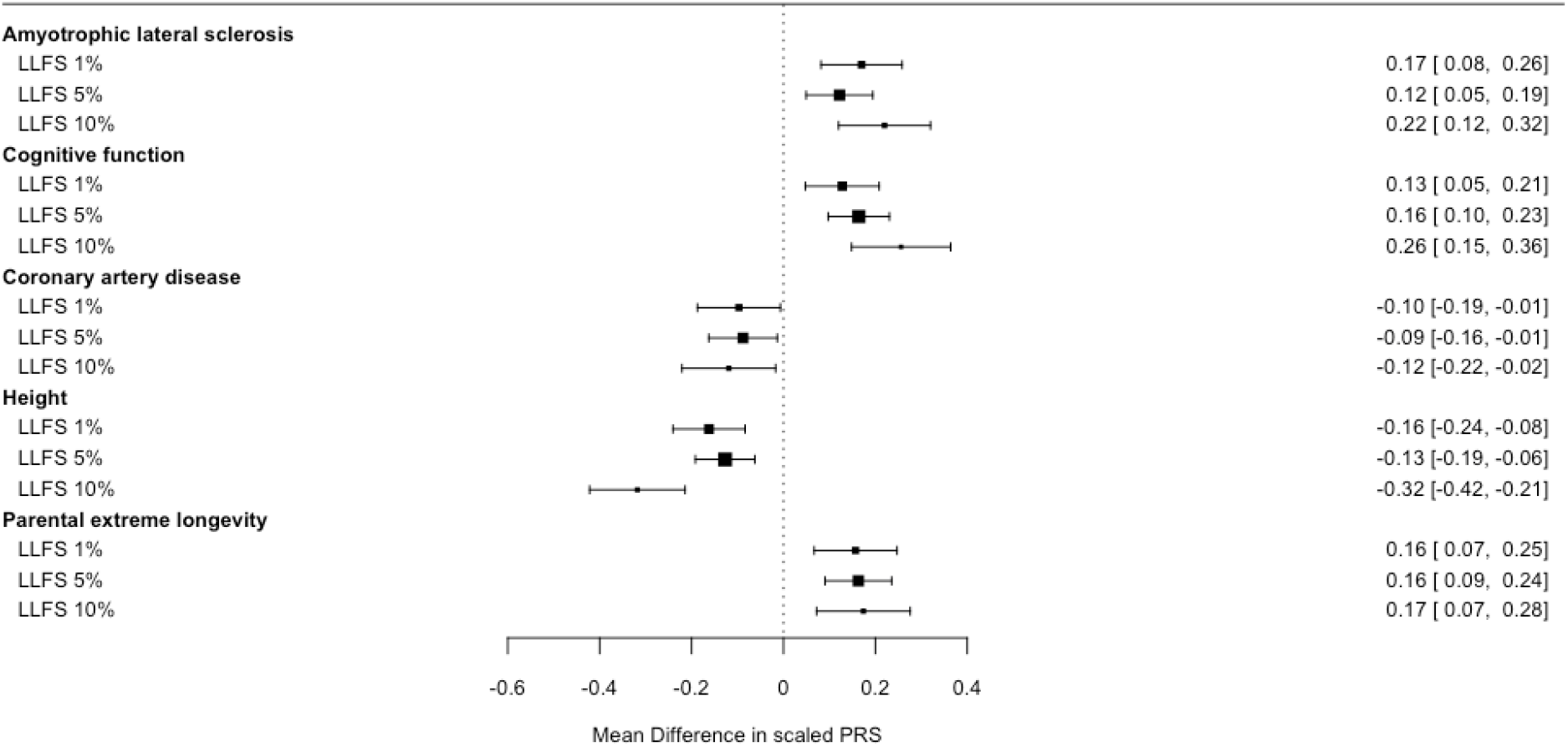
Estimated mean difference and 95% CI of scaled PRS between offspring of ELL individuals and control in LLFS for the traits for which we found a significant mean difference. Extreme longevity of parents was defined by survival thresholds depending on their sex in LLFS. Mean difference is in scaled PRS.

### Association with Disease Risk

We examined the association between AD polygenic risk and incidence of dementia in both NECS and LLFS. This analysis showed that a one standard deviation (SD) increase in AD polygenic risk was associated with a 68% increased hazard for dementia among NECS controls (p = 0.0005), and with more than 100% increased hazard in LLFS controls (p = 0.03), but not in NECS ELLIs (hazard ratio = 1.087, p = 0.18) or top 5% LLFS ELLIs (hazard ratio = 0.991 p = 0.93; Table 1). This pattern persisted even when removing variants in and within a 500KB window of the APOE gene (Table S4).

**Table 1.** Hazard Ratio for CAD and AD PRS and associated diseases

| Study | Score | Outcome | ELLI |  |  | Controls |  |  |
| --- | --- | --- | --- | --- | --- | --- | --- | --- |
|  |  |  | N (N cases) | HR (95% CI) | P value | N (N cases) | HR (95% CI) | P value |
| NECS | AD | Dementia | 977 (263) | 1.087 (0.96, 1.23) | 0.18 | 260 (32) | 1.675 (1.13, 2.48) | 0.0005 |
| LLFS | AD | Dementia | 480 (66) | 0.991 (0.83, 1.19) | 0.93 | 744 (12) | 2.016 (1.08, 3.75) | 0.027 |
| NECS | CAD | CVD | 987 (493) | 1.155 (1.06, 1.26) | 0.001 | 258 (113) | 0.960 (0.78, 1.19) | 0.70 |
| NECS | CAD | Heart Attack | 928 (126) | 1.246 (1.04, 1.49) | 0.0095 | 262 (43) | 0.871 (0.62, 1.23) | 0.43 |
| LLFS | CAD | CVD | 998 (485) | 1.120 (1.02, 1.23) | 0.019 | 738 (138) | 1.411 (1.19, 1.66) | 3.9e-05 |
| LLFS | CAD | Heart Attack | 1031 (112) | 1.335 (1.10, 1.62) | 0.0035 | 747 (28) | 1.327 (0.92, 1.92) | 0.13 |

Study indicates cohort; Score indicates which PRS used; AD for Alzheimer's and CAD for Coronary Artery Disease PRS; Outcome indicates the recorded outcome used for survival analysis; 'N' indicates total number of individuals, while in parentheses 'N cases' indicates number of events in the analysis; HR is per standard deviation increase in PRS; P-value is the corresponding p-value for the PRS in the model predicting time to outcome. Extreme longevity was defined by 5% survival in LLFS and centenarian in NECS. ELL individuals and controls were analyzed separately.

We next assessed the association of the PRS for CAD with incidence of both cardiovascular disease (CVD) and myocardial infarction (MI). Among ELLIs in NECS, a one SD increase in CAD polygenic risk was significantly associated with a 16% increased incidence of CVD (p = 0.001) and a 25% increased incidence of MI (p = 0.0095). Among top 5% ELLIs in LLFS, the PRS for CAD was significantly associated with CVD (HR=1.12, p = 0.02) and MI (HR = 1.34, p = 0.004). In NECS controls, the PRS for CAD score was not significantly associated with MI or CVD, but the number of events was small (n = 113 and 43). In LLFS controls, the PRS for CAD score was significantly associated only with CVD (HR = 1.41, p = 4 × 10^−5^), and not MI.

By contrast, having an AD PRS above the 90^th^ percentile of NECS controls was not associated with dementia in either ELLIs or controls (Table 2). Having a CAD PRS above the 90^th^ percentile of NECS controls was associated with CVD among LLFS controls but not ELLIs, and was not associated with MI among either group.

**Table 2.**
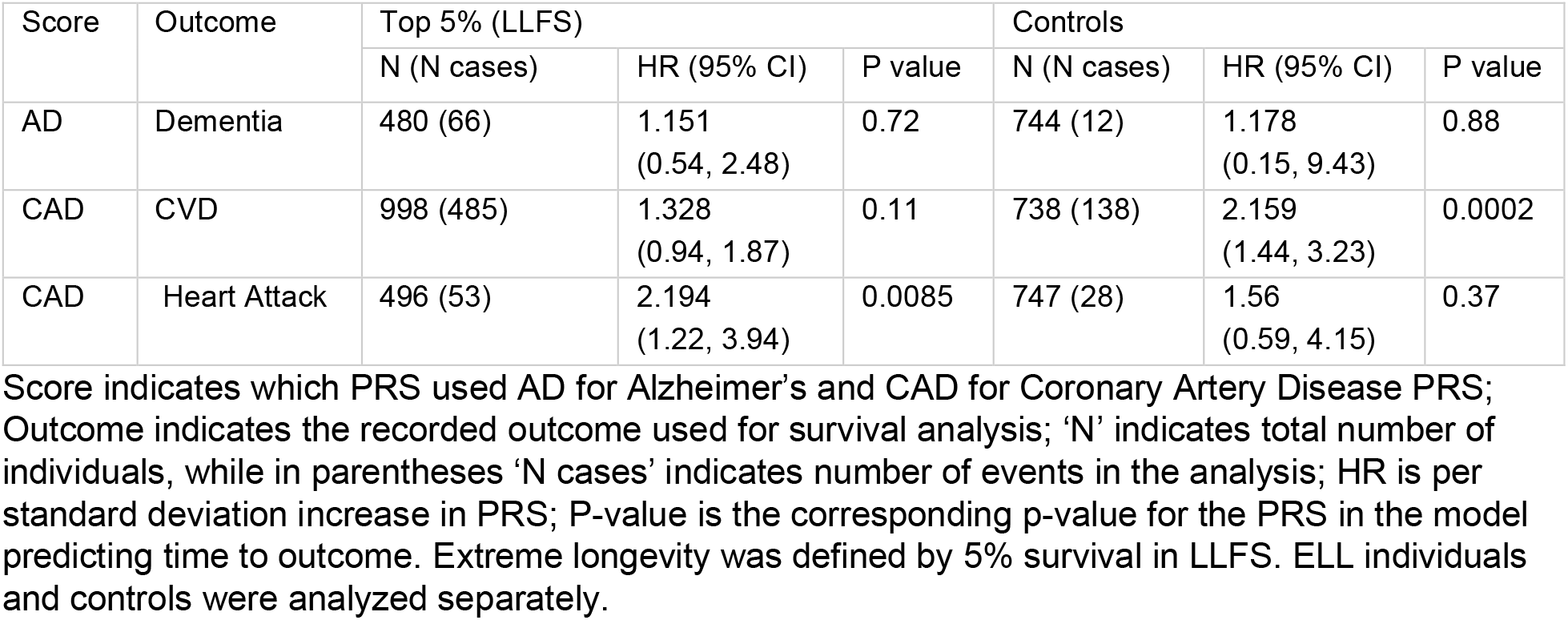
Hazard Ratio for CAD AND AD “high PRS” as defined by the 90% percentile in NECS controls, and associated diseases in LLFS

### Homozygous Scores

We found that ELLIs in both cohorts carried a significantly higher protective homozygous count for both AD and CAD compared to controls (Table 3). In contrast, we did not find a significant mean difference in the risk homozygous count for the same traits, except for risk homozygous count for CAD in NECS. We did detect a significant mean difference in homozygous PRS for both AD and CAD between NECS ELLIs and controls, but only a significant mean difference in homozygous AD PRS in LLFS.

**Table 3.** Mean Difference between ELLI and controls for the three homozygous scores for CAD and AD.

| Study | Base Score | Homozygous Risk count | Homozygous Protective Count | Homozygous PRS (scaled) |
| --- | --- | --- | --- | --- |
| NECS | AD | -0.368 (-0.79, 0.05) | 0.617 (0.25, 1.10) | -0.079 (-0.11, -0.05) |
| NECS | CAD | -1.976 (-2.74, -1.21) | 2.374 (1.66, 3.08) | -0.309 (-0.38, -0.24) |
| LLFS, 5% survival | AD | 0.279 (-0.380, 0.938) | 0.7286 (0.080, 1.377) | -0.1 (-0.233, -0.027) |
| LLFS, 5% survival | CAD | -0.180 (-1.31, 0.950) | 1.316 (0.220, 2.412) | -0.041 (-0.194, 0.006) |

Study indicates cohort; Base Score indicates which PRS used AD for Alzheimer's and CAD for Coronary Artery Disease PRS; Homozygous risk count is the number of homozygous genotypes for risk minor alleles; Homozygous protective count is the number of homozygous genotypes for protective minor alleles; Homozygous PRS is a PRS using the weights from the original score that only assigns genetic risk if an individual is homozygous for the coded allele.

We next associated each of the three types of homozygous scores with incidence of the corresponding disease. We did not find a significant association with any of the scores and either AD or CAD among NECS ELLIs, using an p-value threshold adjusting for three types of our homozygous scores (Table 4). However, we did detect an association between the CAD homozygous PRS with cardiovascular disease among LLFS ELLIs (HR = 1.132, p = 0.009). Among controls, we found an association between the CAD homozygous PRS with cardiovascular disease in LLFS (HR = 1.325, p = 0.003 in NECS), and an association between the AD homozygous PRS and AD protective homozygous count with dementia in NECS (HR = 2.090, p = 0.0002, and HR = 0.887, p = 0.001).

**Table 4.** Hazard Ratio for CAD and AD Homozygous PRSs and associated diseases

| Score | Study | Outcome | ELLI |  | Controls |  |
| --- | --- | --- | --- | --- | --- | --- |
|  |  |  | HR | P value | HR | P value |
| Homozygous PRS CAD | LLFS | CVD | 1.132 (1.031, 1.244) | 0.009 | 1.325 (1.104, 1.591) | 0.0025 |
| Risk count CAD | LLFS | CVD | 1.009 (1.000, 1.018) | 0.042 | 1.012 (0.997, 1.028) | 0.12 |
| Protective count CAD | LLFS | CVD | 0.995 (0.986, 1.004) | 0.24 | 0.980 (0.965, 0.997) | 0.018 |
| Homozygous PRS AD | LLFS | Dementia | 1.045 (0.875, 1.249) | 0.620 | 1.822 (0.928, 3.576) | 0.081 |
| Risk count AD | LLFS | Dementia | 0.997 (0.969, 1.025) | 0.830 | 1.089 (0.985, 1.204) | 0.096 |
| Protective count AD | LLFS | Dementia | 0.992 (0.964, 1.020) | 0.80 | 0.944 (0.856, 1.040) | 0.24 |
| Homozygous PRS CAD | NECS | CVD | 1.096 (1.00, 1.20) | 0.051 | 0.981 (0.819, 1.19) | 0.850 |
| Risk count CAD | NECS | CVD | 1.001 (0.99, 1.01) | 0.74 | 1.001 (0.98, 1.02) | 0.9 |
| Protective count CAD | NECS | CVD | 0.993 (0.98, 1.00) | 0.098 | 1.003 (0.99, 1.02) | 0.74 |
| Homozygous PRS AD | NECS | Dementia | 1.048 (0.93, 1.18) | 0.44 | 2.090 (1.42, 3.08) | 0.00019 |
| Risk count AD | NECS | Dementia | 1.00 (0.98, 1.02) | 0.97 | 1.037 (0.97, 1.11) | 0.31 |
| Protective count AD | NECS | Dementia | 0.996 (0.98, 1.02) | 0.710 | 0.887 (0.83, 0.95) | 0.0013 |

Score indicates which homozygous PRS was used (see table 3 caption for full explanation); Study indicates cohort; Outcome indicates the recorded outcome used for survival analysis; HR is per standard deviation increase in PRS; P-value is the corresponding p-value for the PRS in the model predicting time to outcome. Extreme longevity was defined by 5% survival in LLFS. ELL individuals and controls were analyzed separately. Bonferroni significance threshold adjusted for three types of homozygous scores, $p = 0.05/3 = 0.017$ .

### Parental longevity

In the LLFS, the PRS for parental extreme longevity was a significant predictor of parental longevity for the offspring of top 5% and 1% ELLIs (though not top 10% ELLIs): an increase of one SD in the PRS was associated with a 16% to 19% increased odds of parental longevity (Table 5). However, the PRS for parental extreme longevity was not a significant predictor of parental longevity in the NECS (OR= 1.04, p = 0.56). This lack of significance is likely due to the number of NECS ELLI offspring (N = 487) being much smaller than the number of LLFS ELLI offspring (N = 1213 of top 5%). We estimated an upper threshold of the power of using simple logistic regression to detect an association in NECS to be 0.29, indicating the power of our GEE GLM is ≤ 0.29. This association was even more extreme when the PRS was dichotomized: in the LLFS, having a parental extreme longevity PRS above the 90^th^ percentile in NECS controls increased the odds of parental longevity by 39% to 53% (Table 5).

**Table 5.**
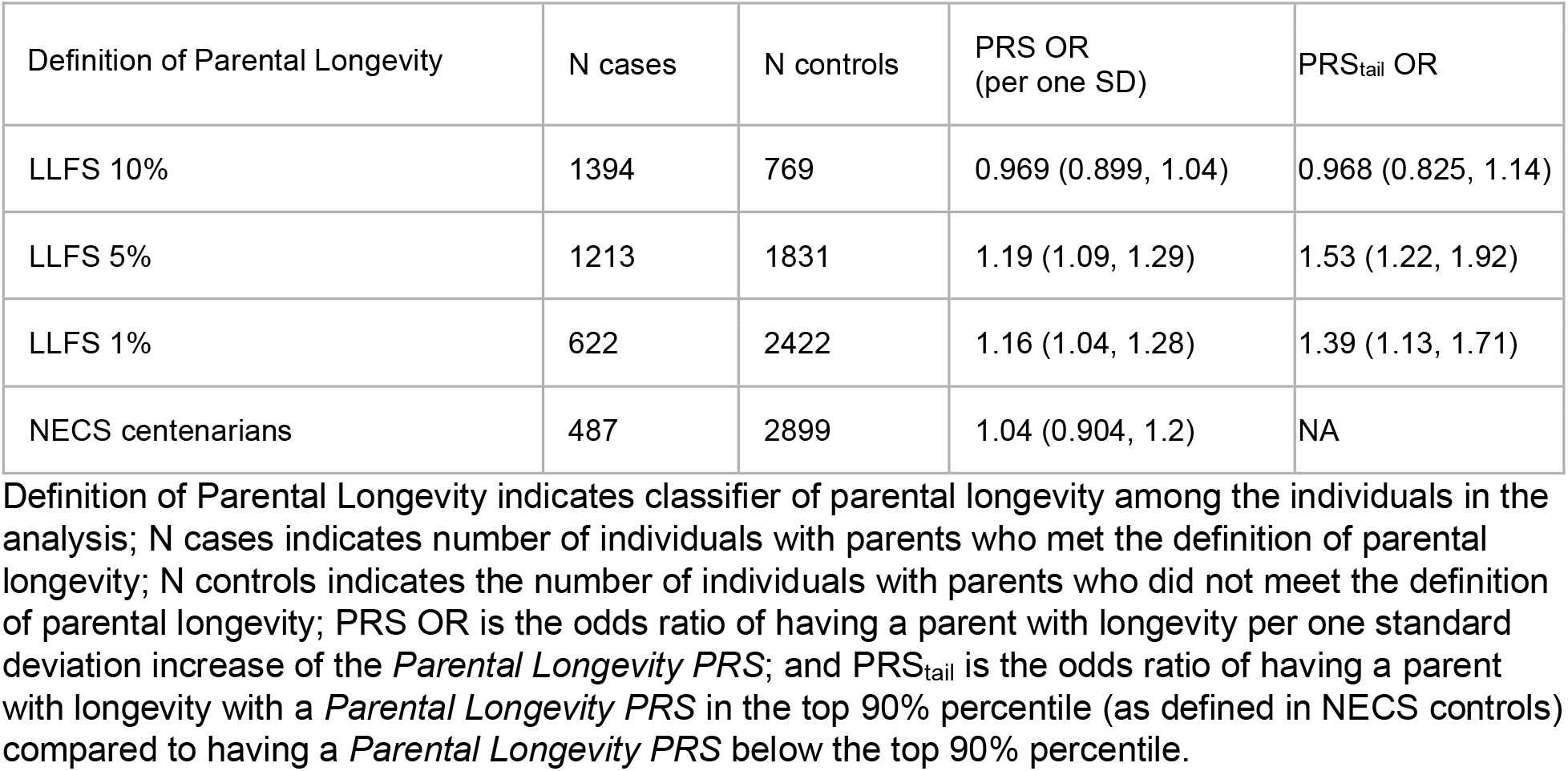
Association of Parental Longevity PRS and parental longevity.

| Definition of Parental Longevity | N cases | N controls | PRS OR<br>(per one SD) | PRS<sub>tail</sub> OR |
| --- | --- | --- | --- | --- |
| LLFS 10% | 1394 | 769 | 0.969 (0.899, 1.04) | 0.968 (0.825, 1.14) |
| LLFS 5% | 1213 | 1831 | 1.19 (1.09, 1.29) | 1.53 (1.22, 1.92) |
| LLFS 1% | 622 | 2422 | 1.16 (1.04, 1.28) | 1.39 (1.13, 1.71) |
| NECS centenarians | 487 | 2899 | 1.04 (0.904, 1.2) | NA |

Definition of Parental Longevity indicates classifier of parental longevity among the individuals in the analysis; N cases indicates number of individuals with parents who met the definition of parental longevity; N controls indicates the number of individuals with parents who did not meet the definition of parental longevity; PRS OR is the odds ratio of having a parent with longevity per one standard deviation increase of the *Parental Longevity PRS*; and PRS<sub>tail</sub> is the odds ratio of having a parent with longevity with a *Parental Longevity PRS* in the top 90% percentile (as defined in NECS controls) compared to having a *Parental Longevity PRS* below the top 90% percentile.

## Discussion

We examined the distribution of 54 PRSs for common and rare complex traits in two cohorts comprising a large number of ELLIs. The PRSs were derived using relaxed thresholds on the level of statistical significance (median of 2979 SNPs, see Table S1). With these comprehensive PRSs, our analysis showed that ELLI are at lower polygenic risk for various aging-related diseases, including in particular AD, and higher polygenic risk for traits related to cognitive function compared to controls. These significant differences in PRS persist at the upper tail of the distributions of the PRSs and with different genetic models (i.e., recessive rather than additive). Thus, our analyses show that ELLIs differ genetically from their normally aging counterparts in polygenic risk. We also demonstrated that these differences are found between ELLI offspring and controls, as expected due to heritability. Further analyzing the ELLI offspring, we were able to replicate the association with polygenic risk for parental longevity found by Pilling et al. [14]. While we know in addition to genetics, environmental and lifestyle factors are shared among families with longevity[22], thus gene-environment interactions may contribute to this relationship, this result provides additional support for the heritability of extreme human longevity.

Our analysis also showed that the PRS for AD was associated with higher risk of dementia in controls but not in ELLIs, while the PRS for CAD was associated with a higher risk of at least one cardiovascular outcome (CVD or MI) in both ELLI and controls. Further, we found an interesting result when considering the association between homozygous genotypes and these two traits of interest. Among ELLIs in both cohorts, the protective homozygous count for AD, and protective homozygous count for CAD was significantly higher than controls, but this was not true of risk homozygous count for the two traits in both cohorts. Among controls in NECS, the protective homozygous count for AD was associated with a reduced hazard of dementia, and among LLFS controls the protective homozygous count for CAD was also associated with *a* reduced hazard of CVD diagnosis. However, these associations were not found in the ELLI. Thus, while we have shown ELLIs have a higher number of protective variants than controls, and that these protective variants protect controls against disease [23], the effect of protective variants do not appear to have measurable effects on incidence of disease in ELLI. Further study into recessive, dominant and additive effects of these variants is needed to understand their impact on longevity. On the whole, these results support two explanations for ELLIs’ resilience to morbidity and mortality: first, that ELLI do in fact have a lower genetic burden of disease-related variants; and second, that whatever genetic burden they do have may be counteracted by (perhaps yet to be discovered) protective genetic and environmental effects [24].

In contrast to previous literature that could not identify a significant difference in the overall burden of disease variants in ELLIs [7, 25], we showed here that, on average, ELLIs carry a lower burden of variants associated with AD, CAD, and lupus. In addition, we showed that ELLIs have higher PRS for cognitive function. Previous work used PRSs defined using a much smaller set of variants; thus, the lack of significant differences in previous analyses may be attributed to poor sensitivity of the PRSs. Our findings differ from recent work [15] that found no significant mean difference in PRSs for various cardiovascular traits between ELLIs and controls. This difference could be due to the lower statistical power of the previous study, which included only 294 centenarians and used a more stringent p-value threshold of 1 × 10^−5^ compared to our 1 × 10^−3^ (thus limiting the number of variants included in the PRSs). Our findings agree with recent observations [14] that parental longevity is associated with lower genetic risk scores for several cardiovascular traits across 75,000 UK biobank participants. Our findings also complement those in [26] finding associations between a lifespan PRS and common diseases in the UKBB. In addition, our results agree with the recent discovery of lower polygenic risk for AD in centenarians and their offspring of Ashkenazi Jewish descent [25]. Our findings regarding AD, CAD and cognitive function are consistent with this previous work, as well as the resilience to AD and CAD seen in ELLIs. Our finding of lower mean PRS for lupus in ELLIs is interesting and more research is necessary to make sense of this result.

It is well known that the prevalence of dementia among ELLIs is low, and those ELLIs who do develop dementia tend to do so very late in life. Our analysis identified a significant reduction in polygenic risk for AD among ELLIs but, surprisingly, higher PRS for AD failed to predict dementia among ELLIs (even omitting the APOE gene & variants in LD (Table S4))[2]. These results suggest that the resilience of ELLIs to dementia may have a genetic component beyond the presence of the protective e2 allele or the absence of the harmful e4 allele in the APOE gene. Discovering these protective variants may lead to therapeutics for healthy cognitive aging.

The large sample size of the LLFS and the large number of ELLIs in the NECS allowed us to examine different definitions of extreme human longevity, using a range of survival probability thresholds. The median age of NECS ELLIs was 103 years, making this cohort ideal for studying exceptional longevity. In contrast, the median age of the older generation in the LLFS was 96, which is admittedly less exceptional. Thus, in our analysis we considered the genetic differences between controls and individuals with both exceptional (i.e. top 1%) and more common (i.e. top 5% and top 10%) longevity. We indeed noticed that some of the PRS differences between ELLIs and controls varied with the longevity threshold used (Figures 1 and 2).

Most of the traits for which we found a significant difference in polygenic risk in one or both studies are important aging-related traits that are significantly less prevalent in ELLIs. However, there were findings in each cohort that surprised us. In NECS, we found that ELLIs had higher mean polygenic risk for ulcerative colitis and inflammatory bowel disease than controls. In LLFS, ELLIs had higher mean polygenic risk than controls for amyotrophic lateral sclerosis (ALS), chronic kidney disease, atopic dermatitis, narcolepsy, psoriasis, gout and primary biliary cholangitis. One explanation for the unexpected finding of higher PRSs for autoimmune traits among ELLIs is that the variants in these PRSs also increase resilience to infection, a common cause of death among the elderly. Indeed, among the genes implicated in ulcerative colitis and inflammatory bowel disease are pro-inflammatory factors like IL22 that are known to be elevated in ELLIs compared to younger individuals [27].The increased mean polygenic risk of ALS found among LLFS ELLIs and their offspring is especially difficult to interpret, though it could be due to some variants in the ALS PRS conferring heterozygote advantage. While previous work has reported a genetic correlation between ALS and late-onset AD [28], we did not find a correlation between the two polygenic scores in either NECS or LLFS, suggesting that the observed association with ALS polygenic risk is not merely due to pleiotropy with AD.

Our work has several limitations. The PRSs were primarily developed from GWAS of individuals of European ancestry, and thus the generalizability of our findings to individuals of other ancestry groups is limited by our study population. We also were only able to include an average of 80.3% of the variants in the original PRS in LLFS and 88.6% in NECS. However, failing to include all the variants should, if anything, lead to more conservative results. Further, we used PRSs that were derived using liberal p-value thresholds to include more genetic variants. While this choice increases the likelihood of including true positives, it also increases the potential for false positives. We do not have reason to believe that this would bias our results, but it is an active area of research. In both study cohorts, medical histories are self-reported, and thus less reliable than medical records. Fortunately, previous work in NECS has indicated that self-reported dementia corresponds well with medical records.

We have demonstrated there are significant differences in polygenic risk for aging-related traits in ELLIs and their offspring, and that for ELLIs, polygenic risk for AD is not predictive of dementia. These findings suggest further work in the genetic differences between ELLIs and controls is necessary. In particular, understanding why polygenic risk is not predictive of dementia in ELLIs may lead to greater insights into both the genetic basis of longevity and AD. We hypothesized that homozygous genotypes could illuminate this result, but all three of our homozygous scores for AD were not associated with dementia in ELLIs. More research is needed to understand what factors buffer the polygenic risk for AD in ELLIs and it is likely that rare variants in centenarians may have a role. In addition, there is much to be explored regarding the gene-gene and gene-environment interactions, as well as epigenetic factors that contribute to the genetics of longevity and AD.

## Data Availability

Data unavailable at this time.

## References

1. Evert, J., et al., Morbidity Profiles of Centenarians. Journal of gerontology, 2003. 58(3): p. 232–237.

2. Andersen, S.L., et al., Health span approximates life span among many supercentenarians: Compression of morbidity at the approximate limit of life span. Journals of Gerontology - Series A Biological Sciences and Medical Sciences, 2012. 67 A(4): p. 395–405.

3. Kheirbek, R.E., et al., Characteristics and Incidence of Chronic Illness in Community-Dwelling Predominantly Male U.S. Veteran Centenarians. Journal of the American Geriatrics Society, 2017. 65(9): p. 2100–2106.

4. Ismail, K., et al., Compression of Morbidity Is Observed Across Cohorts with Exceptional Longevity. Journal of the American Geriatrics Society, 2016. 64(8): p. 1583–1591.

5. Westendorp, R.G.J., et al., Nonagenarian siblings and their offspring display lower risk of mortality and morbidity than sporadic nonagenarians: The Leiden longevity study. Journal of the American Geriatrics Society, 2009. 57(9): p. 1634–1637.

6. Newman, A.B., et al., Health and function of participants in the Long Life family study: A comparison with other cohorts. Aging, 2011. 3(1): p. 63–76.

7. Beekman, M., et al., Genome-wide association study (GWAS)-identified disease risk alleles do not compromise human longevity. Proceedings of the National Academy of Sciences of the United States of America, 2010. 107(42): p. 18046–18049.

8. Manolio, T.A., et al., Finding the missing heritability of complex diseases. Nature, 2009. 461(7265): p. 747–53.

9. Khera, A.V., et al., Genome-wide polygenic scores for common diseases identify individuals with risk equivalent to monogenic mutations. Nat Genet, 2018. 50(9): p. 1219–1224.

10. Stevenson, M., et al., Burden of disease variants in participants of the long life family study. Aging, 2015. 7(2): p. 123–132.

11. Freudenberg-Hua, Y., et al., Disease variants in genomes of 44 centenarians. Mol Genet Genomic Med, 2014. 2(5): p. 438–50.

12. Sebastiani, P. and T.T. Perls, The genetics of extreme longevity: Lessons from the new england centenarian study. Frontiers in Genetics, 2012. 3(NOV): p. 1–7.

13. Sebastiani, P., et al., Genetic signatures of exceptional longevity in humans. PLoS ONE, 2012. 7(1).

14. Pilling, L.C., et al., Human longevity is influenced by many genetic variants: Evidence from 75,000 UK Biobank participants. Aging, 2016. 8(3): p. 547–560.

15. Revelas, M., et al., Exceptional longevity and polygenic risk for cardiovascular health. Genes, 2019. 10(3): p. 1–12.

16. Wainberg, M., et al., Multiomic blood correlates of genetic risk identify presymptomatic disease alterations. Proceedings of the National Academy of Sciences, 2020. 117(35): p. 21813–21820.

17. Young, R.D., et al., Typologies of extreme longevity myths. Curr Gerontol Geriatr Res, 2010. 2010: p. 423087.

18. Sebastiani, P., et al., A family longevity selection score: ranking sibships by their longevity, size, and availability for study. Am J Epidemiol, 2009. 170(12): p. 1555–62.

19. Elo, I.T., et al., Age validation in the long life family study through a linkage to early-life census records. J Gerontol B Psychol Sci Soc Sci, 2013. 68(4): p. 580–5.

20. Sebastiani, P., et al., Genetic signatures of exceptional longevity in humans. PLoS One, 2012. 7(1): p. e29848.

21. Chang, C.C., et al., Second-generation PLINK: rising to the challenge of larger and richer datasets. Gigascience, 2015. 4: p. 7.

22. Perls, T.T., et al., Life-long sustained mortality advantage of siblings of centenarians. Proceedings of the National Academy of Sciences of the United States of America, 2002. 99(12): p. 8442–8447.

23. Giuliani, C., et al., Centenarians as extreme phenotypes: An ecological perspective to get insight into the relationship between the genetics of longevity and age-associated diseases. Mech Ageing Dev, 2017. 165(Pt B): p. 195–201.

24. Ferrucci, L., et al., Measuring biological aging in humans: A quest. Aging Cell, 2020. 19(2): p. e13080.

25. Gutman, D., et al., Similar burden of pathogenic coding variants in exceptionally long-lived individuals and individuals without exceptional longevity. Aging Cell, 2020. 19(10): p. e13216.

26. Timmers, P.R., et al., Genomics of 1 million parent lifespans implicates novel pathways and common diseases and distinguishes survival chances. eLife, 2019. 8: p. 1–40.

27. Basile, G., et al., Healthy centenarians show high levels of circulating interleukin-22 (IL-22). Arch Gerontol Geriatr, 2012. 54(3): p. 459–61.

28. Lu, Q., et al., A Powerful Approach to Estimating Annotation-Stratified Genetic Covariance via GWAS Summary Statistics. Am J Hum Genet, 2017. 101(6): p. 939–964.

