## Supplementary Material for "Distribution of 54 polygenic risk scores for common diseases in long lived individuals and their offspring"

### Supplemental Materials

Table S1 Number of Variants in Original PRS compared to variants computed in the two cohorts

| Outcome | Num. Variants in Original Score | Num. Variants available in LLFS (Prop. of total) | Num. Variants available in NECS (Prop. of total) |
|---|---|---|---|
| Allergic disease | 2979 | 2531 (0.85) | 2788 (0.936) |
| Alzheimer’s disease | 2229 | 1676 (0.752) | 1928 (0.865) |
| Amyotrophic lateral sclerosis | 1989 | 1492 (0.75) | 1711 (0.86) |
| Ankylosing spondylitis | 430 | 405 (0.942) | 414 (0.963) |
| Anxiety tension | 3764 | 2579 (0.685) | 2987 (0.794) |
| Asthma | 2142 | 1769 (0.826) | 1956 (0.913) |
| Atopic dermatitis | 2414 | 1717 (0.711) | 1941 (0.804) |
| Atrial fibrillation | 5148 | 3709 (0.72) | 4172 (0.81) |
| Bipolar disorder | 1957 | 1485 (0.759) | 1690 (0.864) |
| Birth weight | 4114 | 2916 (0.709) | 3343 (0.813) |
| Body mass index | 10825 | 10500 (0.97) | 10701 (0.989) |
| Breast cancer | 5731 | 4278 (0.746) | 4841 (0.845) |
| Carpal tunnel syndrome | 2262 | 1891 (0.836) | 2085 (0.922) |
| Celiac disease | 714 | 677 (0.948) | 696 (0.975) |
| Chronic kidney disease | 640 | 610 (0.953) | 625 (0.977) |
| Chronotype | 7877 | 6111 (0.776) | 6783 (0.861) |
| Cognitive performance | 7515 | 6016 (0.801) | 6612 (0.88) |
| Coronary artery disease | 4693 | 3967 (0.845) | 4357 (0.928) |
| Crohn’s disease | 5557 | 3804 (0.685) | 4510 (0.812) |
| Depression | 4389 | 3754 (0.855) | 4126 (0.94) |
| Diastolic blood pressure | 3705 | 2569 (0.693) | 2915 (0.787) |
| Educational attainment | 4070 | 3652 (0.897) | 3880 (0.953) |
| Epilepsy | 1863 | 1852 (0.994) | 1855 (0.996) |
| FEV1 | 8974 | 7074 (0.788) | 7846 (0.874) |
| Glaucoma | 2599 | 2296 (0.883) | 2457 (0.945) |
| Gout | 918 | 787 (0.857) | 842 (0.917) |
| Heel bone mineral density | 20137 | 14588 (0.724) | 16907 (0.84) |
| Height | 20610 | 19649 (0.953) | 20259 (0.983) |
| Inflammatory bowel disease | 6501 | 4405 (0.678) | 5188 (0.798) |
| Insomnia symptoms | 5005 | 3650 (0.729) | 4105 (0.82) |
| Intelligence | 7536 | 6541 (0.868) | 7035 (0.934) |
| Juvenile idiopathic arthritis | 181 | 179 (0.989) | 180 (0.994) |
| Life satisfaction | 638 | 631 (0.989) | 635 (0.995) |
| Male pattern baldness | 10437 | 7842 (0.751) | 8884 (0.851) |
| Multiple sclerosis | 576 | 546 (0.948) | 562 (0.976) |
| Narcolepsy | 130 | 118 (0.908) | 121 (0.931) |
| Neuroticism | 3144 | 3108 (0.989) | 3132 (0.996) |
| Parental extreme longevity | 1913 | 1398 (0.731) | 1622 (0.848) |
| Primary biliary cholangitis | 813 | 799 (0.983) | 799 (0.983) |
| Primary sclerosing cholangitis | 1683 | 1524 (0.906) | 1596 (0.948) |
| Prostate cancer | 3871 | 2696 (0.696) | 3128 (0.808) |
| Psoriasis | 585 | 538 (0.92) | 561 (0.959) |
| Rheumatoid arthritis | 2339 | 2068 (0.884) | 2232 (0.954) |
| Sleep duration | 5449 | 4058 (0.745) | 4550 (0.835) |
| Stroke | 2383 | 2118 (0.889) | 2281 (0.957) |
| Subjective well being | 964 | 934 (0.969) | 958 (0.994) |

| | | | |
|---|---|---|---|
| Systemic lupus erythematosus | 2801 | 2408 (0.86) | 2617 (0.934) |
| Systolic blood pressure | 3661 | 2603 (0.711) | 2931 (0.801) |
| Total body bone mineral density | 3993 | 2922 (0.732) | 3339 (0.836) |
| Type 1 diabetes | 444 | 407 (0.917) | 428 (0.964) |
| Type 2 diabetes | 399 | 381 (0.955) | 389 (0.975) |
| Ulcerative colitis | 4927 | 3362 (0.682) | 3943 (0.8) |
| WHRadjBMI | 317 | 292 (0.921) | 302 (0.953) |
| Worry | 4857 | 4122 (0.849) | 4498 (0.926) |

Number of Variants in Original Score compared to number of variants computed in LLFS and NECS cohorts. Average number of variants computed per score in LLFS is 80.6 and in NECS is 88.6.

Table S2a-c. Description of Study Participants by Study

Table S2a. Study participants by definition of ELL in NECS

| Definition of Longevity | N | Median Age & Range | Proportion Female |
|---|---|---|---|
| Centenarians | 1495 | 103 (88 – 119) | 0.73 |
| Controls | 2899 | 82 (55 – 100) | 0.63 |
| Offspring | 487 | 85 (55 – 102) | 0.59 |

Table S2b. Study participants by definition of ELL in LLFS

| Definition of Longevity | N | Median Age & Range | Proportion Female |
|---|---|---|---|
| 1% survival threshold | 502 | 101 (97 – 111) | 0.41 |
| 5% survival threshold | 1044 | 98 (91 – 111) | 0.43 |
| 10% survival threshold | 1270 | 97 (87 – 110) | 0.47 |
| Controls | 761 | 72 (27 – 97) | 0.45 |

Table S2c. Study participants by definition of Parental Longevity in LLFS

| Definition of parental longevity | N | Median Age & Range |
|---|---|---|
| 1% survival threshold | 622 | 73 (50 – 93) |
| 5% survival threshold | 1213 | 72 (46 – 94) |
| 10% survival threshold | 1394 | 71 (46 – 93) |
| Controls | 748 | 72 (27 – 97) |

Table S3. Estimated mean difference and 95% CI of scaled PRS between ELLI and controls for all traits studied

| **PRS** | **NECS** | **LLFS 1%** | **LLFS 5%** | **LLFS 10%** |
|---|---|---|---|---|
| Allergic disease PRS | 0.013 (-0.054, 0.08) | -0.038 (-0.158, 0.082) | -0.034 (-0.138, 0.07) | -0.039 (-0.139, 0.061) |
| Alzheimer's disease PRS | -0.291 (-0.358, -0.224) | -0.211 (-0.334, -0.088) | -0.218 (-0.323, -0.113) | -0.205 (-0.306, -0.104) |
| Amyotrophic lateral sclerosis PRS | -0.031 (-0.096, 0.034) | 0.353 (0.233, 0.473) | 0.277 (0.173, 0.38) | 0.281 (0.181, 0.38) |
| Ankylosing spondylitis PRS | 0.001 (-0.052, 0.054) | -0.102 (-0.223, 0.019) | -0.087 (-0.191, 0.017) | -0.091 (-0.191, 0.009) |
| Anxiety tension PRS | 0.038 (-0.029, 0.105) | 0.041 (-0.083, 0.164) | 0.023 (-0.081, 0.127) | 0.014 (-0.087, 0.115) |

| | | | | |
|---|---|---|---|---|
| Asthma PRS | -0.052 (-0.12, 0.015) | 0.103 (-0.017, 0.222) | 0.122 (0.018, 0.226) | 0.112 (0.011, 0.212) |
| Atopic dermatitis PRS | -0.02 (-0.084, 0.045) | 0.288 (0.167, 0.41) | 0.229 (0.125, 0.333) | 0.229 (0.129, 0.329) |
| Atrial fibrillation PRS | -0.123 (-0.19, -0.055) | -0.145 (-0.266, -0.023) | -0.119 (-0.223, -0.016) | -0.105 (-0.205, -0.005) |
| Bipolar disorder PRS | 0.021 (-0.046, 0.089) | 0.134 (0.013, 0.255) | 0.112 (0.008, 0.216) | 0.097 (-0.003, 0.197) |
| Birth weight PRS | -0.004 (-0.068, 0.059) | -0.218 (-0.339, -0.098) | -0.171 (-0.273, -0.068) | -0.183 (-0.283, -0.083) |
| Body mass index PRS | -0.285 (-0.352, -0.218) | -0.134 (-0.252, -0.016) | -0.095 (-0.197, 0.006) | -0.088 (-0.187, 0.011) |
| Breast cancer PRS | -0.053 (-0.117, 0.011) | 0.155 (0.034, 0.275) | 0.182 (0.079, 0.286) | 0.196 (0.096, 0.296) |
| Carpal tunnel syndrome PRS | -0.02 (-0.085, 0.045) | 0.025 (-0.097, 0.147) | 0.002 (-0.102, 0.105) | 0 (-0.101, 0.1) |
| Celiac disease PRS | -0.01 (-0.039, 0.019) | -0.118 (-0.24, 0.004) | -0.112 (-0.215, -0.008) | -0.1 (-0.201, 0.001) |
| Chronic kidney disease PRS | -0.03 (-0.089, 0.03) | 0.302 (0.183, 0.422) | 0.258 (0.154, 0.362) | 0.266 (0.165, 0.367) |
| Chronotype PRS | -0.04 (-0.108, 0.027) | -0.187 (-0.309, -0.064) | -0.148 (-0.251, -0.046) | -0.168 (-0.268, -0.069) |
| Cognitive function PRS | 0.104 (0.048, 0.16) | 0.288 (0.169, 0.407) | 0.324 (0.22, 0.428) | 0.314 (0.213, 0.414) |
| Cognitive performance PRS | 0.088 (0.023, 0.154) | 0.057 (-0.063, 0.177) | 0.093 (-0.011, 0.197) | 0.091 (-0.009, 0.192) |
| Coronary artery disease PRS | -0.34 (-0.407, -0.274) | -0.241 (-0.364, -0.119) | -0.245 (-0.348, -0.143) | -0.252 (-0.351, -0.152) |
| Crohn’s disease PRS | 0.047 (-0.018, 0.112) | 0.176 (0.056, 0.297) | 0.157 (0.054, 0.259) | 0.165 (0.065, 0.265) |
| Depression PRS | -0.054 (-0.121, 0.012) | 0.017 (-0.104, 0.139) | 0.046 (-0.058, 0.15) | 0.019 (-0.081, 0.12) |
| Diastolic blood pressure PRS | -0.083 (-0.149, -0.016) | -0.084 (-0.205, 0.037) | -0.177 (-0.281, -0.073) | -0.189 (-0.289, -0.088) |
| Educational attainment PRS | 0.115 (0.052, 0.179) | 0.155 (0.037, 0.273) | 0.189 (0.084, 0.293) | 0.19 (0.09, 0.291) |
| Epilepsy PRS | 0.048 (-0.019, 0.115) | -0.017 (-0.139, 0.105) | -0.013 (-0.117, 0.092) | -0.026 (-0.127, 0.074) |
| FEV1 PRS | -0.001 (-0.058, 0.056) | 0.173 (0.05, 0.295) | 0.153 (0.049, 0.258) | 0.143 (0.043, 0.243) |
| Glaucoma PRS | -0.034 (-0.102, 0.034) | 0.063 (-0.054, 0.179) | 0.051 (-0.052, 0.154) | 0.024 (-0.076, 0.123) |
| Gout PRS | -0.009 (-0.07, 0.052) | 0.219 (0.099, 0.34) | 0.226 (0.121, 0.33) | 0.215 (0.115, 0.315) |
| Heel bone mineral density PRS | -0.1 (-0.167, -0.032) | -0.021 (-0.139, 0.097) | -0.041 (-0.144, 0.062) | -0.044 (-0.144, 0.056) |
| Height PRS | -0.018 (-0.075, 0.04) | -0.435 (-0.553, -0.317) | -0.437 (-0.54, -0.334) | -0.428 (-0.527, -0.329) |
| Inflammatory bowel disease PRS | 0.105 (0.039, 0.17) | 0.091 (-0.033, 0.216) | 0.086 (-0.019, 0.19) | 0.094 (-0.008, 0.196) |
| Insomnia symptoms PRS | -0.082 (-0.15, -0.014) | 0.109 (-0.012, 0.23) | 0.077 (-0.026, 0.18) | 0.057 (-0.042, 0.156) |
| Juvenile idiopathic arthritis PRS | -0.048 (-0.112, 0.017) | 0.019 (-0.104, 0.142) | 0.058 (-0.046, 0.162) | 0.05 (-0.051, 0.15) |
| Life satisfaction PRS | 0.027 (-0.038, 0.092) | 0.091 (-0.032, 0.213) | 0.038 (-0.066, 0.142) | 0.027 (-0.074, 0.128) |
| Male pattern baldness PRS | 0 (-0.068, 0.067) | -0.12 (-0.242, 0.003) | -0.115 (-0.218, -0.012) | -0.103 (-0.203, -0.003) |
| Multiple sclerosis PRS | 0.025 (-0.017, 0.068) | -0.173 (-0.293, -0.053) | -0.161 (-0.264, -0.058) | -0.125 (-0.225, -0.025) |
| Narcolepsy PRS | -0.061 (-0.125, 0.003) | 0.245 (0.124, 0.366) | 0.199 (0.094, 0.304) | 0.203 (0.102, 0.304) |
| Neuroticism PRS | 0.009 (-0.058, 0.076) | 0.002 (-0.119, 0.124) | -0.011 (-0.115, 0.093) | -0.007 (-0.107, 0.094) |
| Parental extreme longevity PRS | 0.093 (0.029, 0.158) | 0.21 (0.092, 0.328) | 0.253 (0.15, 0.356) | 0.226 (0.126, 0.325) |
| Primary biliary cholangitis PRS | 0.013 (-0.044, 0.07) | 0.212 (0.091, 0.333) | 0.197 (0.093, 0.3) | 0.186 (0.086, 0.286) |
| Primary sclerosing cholangitis PRS | -0.011 (-0.045, 0.023) | -0.247 (-0.368, -0.126) | -0.209 (-0.311, -0.107) | -0.182 (-0.282, -0.083) |
| Prostate cancer PRS | -0.07 (-0.135, -0.004) | -0.25 (-0.371, -0.129) | -0.267 (-0.371, -0.163) | -0.264 (-0.364, -0.164) |
| Psoriasis PRS | 0.009 (-0.057, 0.074) | 0.226 (0.108, 0.344) | 0.217 (0.115, 0.319) | 0.219 (0.12, 0.319) |
| Rheumatoid arthritis PRS | -0.008 (-0.061, 0.046) | -0.006 (-0.129, 0.117) | 0.031 (-0.073, 0.134) | -0.004 (-0.105, 0.096) |
| Sleep duration PRS | 0.018 (-0.049, 0.084) | -0.126 (-0.245, -0.007) | -0.143 (-0.246, -0.041) | -0.152 (-0.251, -0.052) |
| Stroke PRS | -0.227 (-0.294, -0.16) | -0.132 (-0.252, -0.012) | -0.136 (-0.239, -0.033) | -0.119 (-0.219, -0.02) |
| Subjective well-being PRS | -0.051 (-0.118, 0.017) | -0.04 (-0.159, 0.079) | 0.016 (-0.088, 0.119) | 0.032 (-0.068, 0.132) |
| Systemic lupus | -0.07 (-0.111, -0.03) | -0.202 (-0.324, -0.079) | -0.178 (-0.282, -0.074) | -0.174 (-0.275, -0.073) |

| | | | | |
|---|---|---|---|---|
| erythematosus PRS | | | | |
| Systolic blood pressure PRS | -0.167 (-0.235, -0.099) | -0.013 (-0.134, 0.108) | -0.089 (-0.193, 0.015) | -0.092 (-0.192, 0.008) |
| Total body bone mineral density PRS | 0.027 (-0.041, 0.095) | 0.07 (-0.05, 0.19) | -0.019 (-0.122, 0.085) | -0.019 (-0.119, 0.08) |
| Type 1 diabetes PRS | -0.086 (-0.153, -0.02) | 0.068 (-0.053, 0.188) | 0.093 (-0.01, 0.196) | 0.089 (-0.011, 0.189) |
| Type 2 diabetes PRS | -0.082 (-0.148, -0.015) | 0.125 (0.006, 0.245) | 0.089 (-0.014, 0.191) | 0.084 (-0.015, 0.184) |
| Ulcerative colitis PRS | 0.118 (0.053, 0.183) | -0.143 (-0.263, -0.023) | -0.063 (-0.167, 0.04) | -0.047 (-0.147, 0.053) |
| WHRadjBMI PRS | -0.055 (-0.122, 0.011) | -0.159 (-0.281, -0.036) | -0.199 (-0.303, -0.095) | -0.183 (-0.284, -0.083) |
| Worry PRS | 0.003 (-0.063, 0.069) | 0.039 (-0.085, 0.163) | 0.011 (-0.095, 0.117) | 0.003 (-0.099, 0.104) |

Extreme longevity was defined by survival thresholds depending on their sex in LLFS and centenarian status in NECS. Mean difference is in scaled PRS. While the GWAS used for our Cognitive Function PRS originally labelled the trait “Intelligence”, the study was a meta-analysis of two studies, one that used Spearman’s g as the outcome, and one that used college entrance exams taken by the subjects at 12 years of age. We believe the label of “Intelligence” is far too broad to describe the actual phenotype used in these studies, and have therefore chosen to use the label “Cognitive Function” in its stead.

Table S4a-b. Results from removing APOE and surrounding variants from Alzheimer’s PRS

Table S4a. Mean difference in scaled PRS between ELLI and controls omitting APOE variants and all variants within 500kb

| Definition of ELLI | Mean Difference in Scaled PRS (95% CI) |
|---|---|
| NECS | -0.149 (-0.217, -0.081) |
| LLFS 1% | -0.010 (-0.130, 0.110) |
| LLFS 5% | -0.0353 (-0.139, 0.068) |
| LLFS 10% | 0.035 (-0.135, 0.065) |

Table S4b. Hazard Ratio for PRS omitting APOE variants and all variants within 500kb and onset of dementia

| Study | ELLI | | | Controls | | |
|---|---|---|---|---|---|---|
| | N (N cases) | HR (95% CI) | P value | N (N cases) | HR (95% CI) | P value |
| NECS | 977 (263) | 0.996 (0.88, 1.12) | 0.94 | 260 (32) | 1.556 (1.04, 2.34) | 0.032 |
| LLFS | 480 (66) | 1.003 (0.84, 1.20) | 0.97 | 744 (12) | 2.136 (1.06, 4.30) | 0.033 |

Study indicates cohort; ‘N’ indicates total number of individuals, while in parentheses ‘N cases’ indicates number of events in the analysis; HR is per standard deviation increase in PRS and 95% CI; P-value is the corresponding p-value for the PRS in the model predicting time to dementia. Extreme longevity was defined by 5% survival in LLFS. ELLI and controls were analyzed separately.

Additional Results

Among the controls, we found a significant association between homozygous PRS for CAD and the number of protective homozygous genotypes with cardiovascular disease in the LLFS (HR=1.325, p=0.0025, and HR=0.98, p=0.018 respectively). Further, we found a significant association between homozygous PRS for Alzheimer’s and the number of protective homozygous genotypes with Dementia in NECS controls (HR = 2.09, p=0.00019 and HR=0.887, p=0.0013respectively).
